# Pilot study on the use of low molecular weight heparins in the prevention of thromboembolic disease during pregnancy and puerperium

**DOI:** 10.1101/2022.04.26.22274264

**Authors:** Paloma Anchústegui-Mendizábal, Patricia Anchústegui-Mendizábal, Laura Arechabala-Palacios, Laura Fernández-González, Clara Garcia-Gil, Jesús Miguel Hernández-Guijo, Óscar Martínez-Pérez

**Affiliations:** Universidad Autonoma de Madrid.Spain; Hospital Universitario Puerta de Hierro.Majadahonda.Spain

**Keywords:** pregnancy, venous thromboembolism prophylaxis, low molecular weight heparin, risk factors, risk assessment, *SARS-CoV-2*

## Abstract

A pregnant woman is 4 to 5 times more likely to suffer a thromboembolic event than a non-pregnant woman. Furthermore, an increase in these episodes has been observed in women infected with *SARS-CoV-2*. Consequently, the prophylactic prescription of low-molecular-weight heparins (LMWH) in pregnant women is undergoing an increase that has not been evaluated yet. The aim of this study was to determine the prevalence of LMWH prescription in pregnant women at the *Hospital Universitario Puerta de Hierro Majadahonda* (HUPHM), according to their level of risk and its variation due to *SARS-CoV-2* infection. To answer this question, a retrospective cohort of 113 women who gave birth during the month of February at the HUPHM was designed. The level of individual risk of thromboembolism, according to the Royal College guidelines (37a), was calculated with an interview to complete a questionnaire and the analysis of their clinical records. 53.6% of the women were prescribed LMWH as indicated in the guidelines. This high prevalence is explained by the high age of the pregnant women (over 35 years), the wave of the omicron variant (December 2021) and a high rate of cesarean sections (25%). On the other hand, the percentage of patients with COVID-19 was 17.6% but only 53% of them had received LMWH. In conclusion, LMWH is a very common prescription in obstetrics, mostly during puerperium, and has become even more relevant due to the COVID-19 pandemic

## Introduction

In Western societies, the increase in life expectancy, changes in values, as well as the new assisted reproduction techniques have considerably delayed the age of having the first child. A pregnant woman is 4 to 5 times more likely to suffer a thromboembolic event than a non-pregnant one, with venous thromboembolism being one of the leading causes of maternal death worldwide (1). Due to the growing number of pregnant women with risk factors, the incidence of venous thromboembolic disease (VTE) has increased in recent decades (2).

The mortality and morbidity of VTE are potentially preventable, as more than two-thirds of these women have identifiable risk factors and can benefit from adequate thromboprophylaxis (3). The likelihood is even greater if the *SARS-CoV-2* infection is added. The current pandemic of COVID-19 has evidenced a high incidence of thromboembolic events in patients with the infection, fatal in many cases. In addition, there is a lack of knowledge about the incidence of heparin use and recommendations about its use during pregnancy and *SARS-CoV-2* infection. To date, there are no studies published or under development in Spain that analyze the prevalence during pregnancy of risk factors for VTE in pregnant women and the administration of heparins.

### Hypotheses and objectives

There is a lack of knowledge about the distribution of VTE risk factors in the obstetric population, aggravated by the influence of the pandemic. Our hypothesis suggests that the prevalence of heparin prescription during pregnancy is high and has been increased due to the COVID-19 pandemic.

Consequently, the primary objective of this pilot study is to determine the prevalence of prescription of low molecular weight heparins (LMWH) in pregnant women, according to their level of risk for the development of VTE, as well as the variation of this risk due to *SARS-CoV-2* infection. As secondary objectives, we sought to clarify whether LMWH prophylaxis was carried out according to the risk level of each patient and whether it conforms to the indications of the Royal College guidelines for the prophylactic treatment of VTE.

## Materials and methods

The pilot study protocol was designed based on the variables of the ETV-OBS risk calculator (4). The design was based on a literature search in Pubmed on the use of antithrombotics in pregnancy and the Royal College guidelines (37a) on thromboembolic risk in pregnant women were studied.

A database was created by completing 3 questionnaires: first trimester (before week 14 of gestation), third trimester (between week 24 and 26) and postnatal; together with the analysis of medical records.

After this data collection, the results obtained were analyzed, comparing in each of the three periods analyzed the risk level of each patient and whether or not LMWH prophylaxis was carried out, according to the indications of the Royal College guidelines.

### Variables

The database was constructed based on the variables presented below and measured in each patient.

The variables measured in the 1st and 3rd trimester were: age, weight (kg), height (m), body mass index (BMI), number of cigarettes/day, gravidity (number of pregnancies), parity (number of living children), family history of venous thromboembolic disease, personal history of venous thromboembolic disease, high risk thrombophilia, low risk thrombophilia without family history and low risk thrombophilia with family history, important medical comorbidities of the patient, varicose veins, long-term travel, immobility, hospitalization, surgery, infection, preeclampsia, multiple pregnancy, pregnancy after *in vitro* fertilization (IVF), hyperemesis or dehydration, type of heparin if administered, heparin dose and reason for administration, treatment with acetylsalicylic acid (ASA), dose and reason for administration.

The variables measured during postpartum were: all of the above except for pregnancy. Also added: gestational age (gestational week at birth), type of delivery (euthyroid, planned or emergency cesarean section and instrumental delivery), 24 hours or longer delivery, postpartum hemorrhage, abortion, preterm delivery (less than 37 weeks).

### Inclusion and exclusion criteria

The inclusion criteria followed were: that the patients were over age, that they signed the informed consent form, that they had given birth, and that they were admitted to the gynecology and obstetrics department of the *Hospital Universitario Puerta de Hierro Majadahonda* (HUPHM). We went to this hospital during the month of February 2022. If one or more of the above criteria were not met, the patient was excluded from the study.

### Ethical considerations

Prior to the start of this study, the members of the study took the courses offered by The Global Health Network: “Introduction to Clinical Research” and “Good Clinical Practice Guidelines ICH E6 (R2) “.

The trial was conducted in accordance with Good Clinical Practice guidelines and was sent for assessment by the Bioethics Committee of the HUPHM. The participants were informed of the characteristics and objectives of the study, as well as the confidentiality and protection of the data to be collected, and written informed consent was obtained from all the participants. The method of anonymization followed was the assignment of a consecutive number to each patient.

The data were collected by the investigators, analyzed and interpreted by the same. The authors are responsible for the accuracy and completeness of the data as well as for the fidelity of the trial to the protocol.

Of the patients admitted, 15 patients did not meet the inclusion criteria as they were hospitalized for reasons other than given birth. The 113 women who met the inclusion criteria were offered informed consent. Five of them refused consent, while 108 gave consent (see Figure 1).

**Figure 1.**
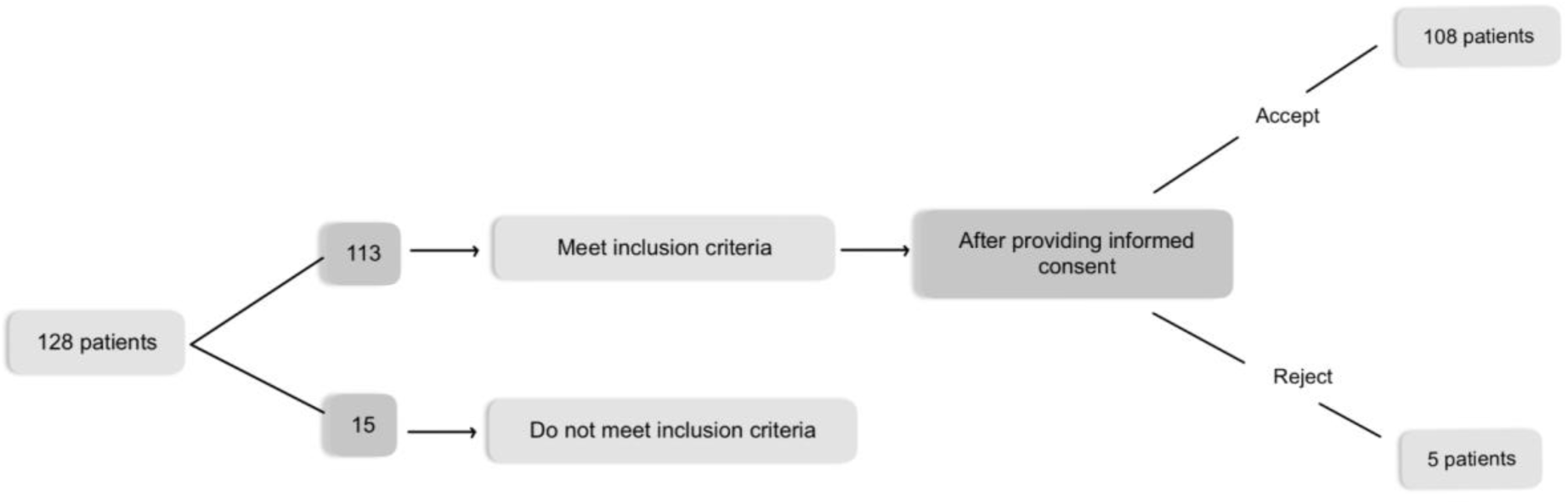
Flow diagram. Diagram showing the flow of patients throughout the study.

## Results

### Demographics of the series

Figure 2 summarizes the demographic characteristics of the 108 patients included in the study at the first obstetric visit that could impact the baseline risk for VTE in pregnant women. Of the 108 women, 38% were older than 35 years, 31.5% of the total were between 36 and 40 years of age and 6.5% between 41 and 45 years. Only 1.9% were between 21 and 25 years of age, 15.7% were in the 26-30 age range and, the vast majority of the series, 44.4% were between 31 and 35 years of age, both included. Of all women, 61.1% had normal weight, 7.4% were obese and 22.2% were overweight compared to 9.3% who were underweight. Of the patients studied, 11.1% were smokers during pregnancy. The percentage of primigravidae women was 39.8%: 50% of women aged 21-25 years, 64.7% of women aged 26-30 years, 37.5% of women aged 31-35 years, 29.4% of women aged 36-40 years and 42.9% of women aged 41-45 years. The numerical differences between pregnancy and parity are due to abortions.

**Figure 2.**
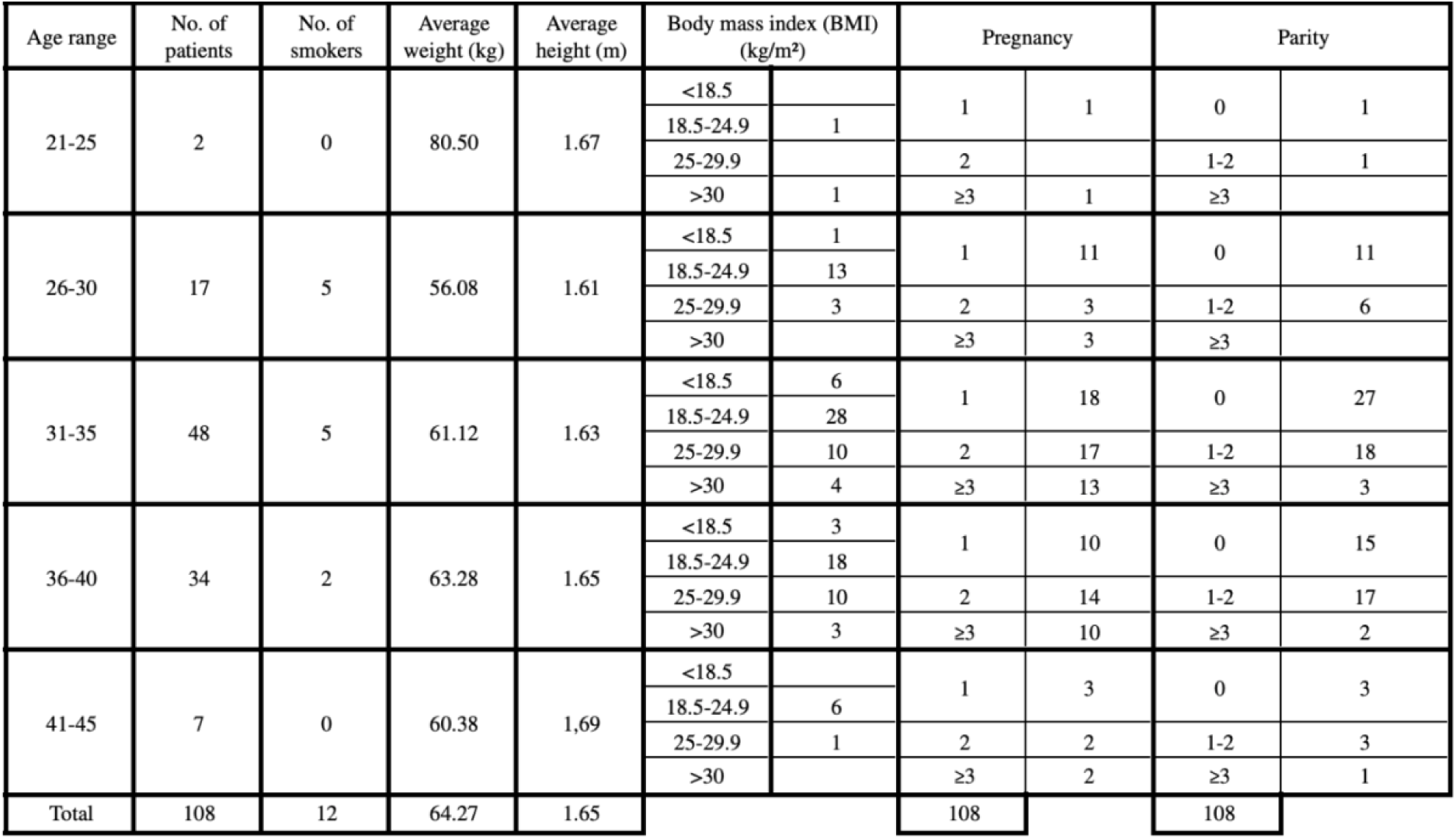
Demographics of the series. It is grouped by 5-year age ranges. Weight is measured in kilograms, height in meters and BMI is grouped into 4 ranges (corresponding to underweight, normal weight, overweight and obesity). Gravidity (pregnancy) is determined in ranges of 1, 2 and greater than or equal to 3 and parity in ranges of 0, 1 or 2 and greater than or equal to 3.

### History related to VTE

After the demographic analysis, the VTE-related history of the patients in the series was studied (Figure 3). None had a personal history of previous VTE.

**Figure 3.**
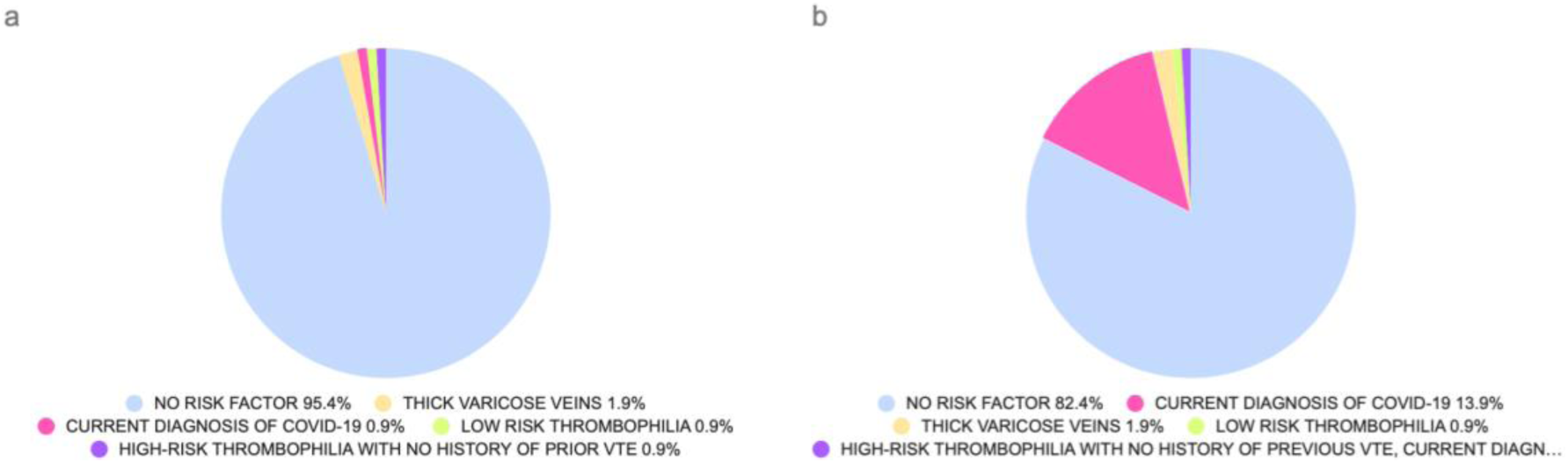
Risk factors in personal history. Pie chart comparing personal history as a risk factor (indicated in the legend) in the first (a) and third trimester of gestation (b).

However, there was a family history of VTE in 1.8% of the patients. Finally, we counted the number of patients who had been immobilized during pregnancy, and the result was 1.8% in the first trimester, corresponding to women who had taken a long trip. In the third trimester, none of the patients had suffered immobilization.

Of note was the increase in the number of women diagnosed with *SARS-CoV-2* infection in the third trimester (13.9%) compared to the first trimester of gestation (0.9%).

### Comparison of the first and third trimester of gestation

Figure 4 compares the risk factors between the first and third trimester of gestation (excluding personal history). The number of cigarettes consumed per day did not vary during this period. The number of patients with antenatal risk factors increased from 16 to 20 from the first to the third trimester, and this variation is explained by the increase in pregnancies with risk of preeclampsia.

**Figure 4.**
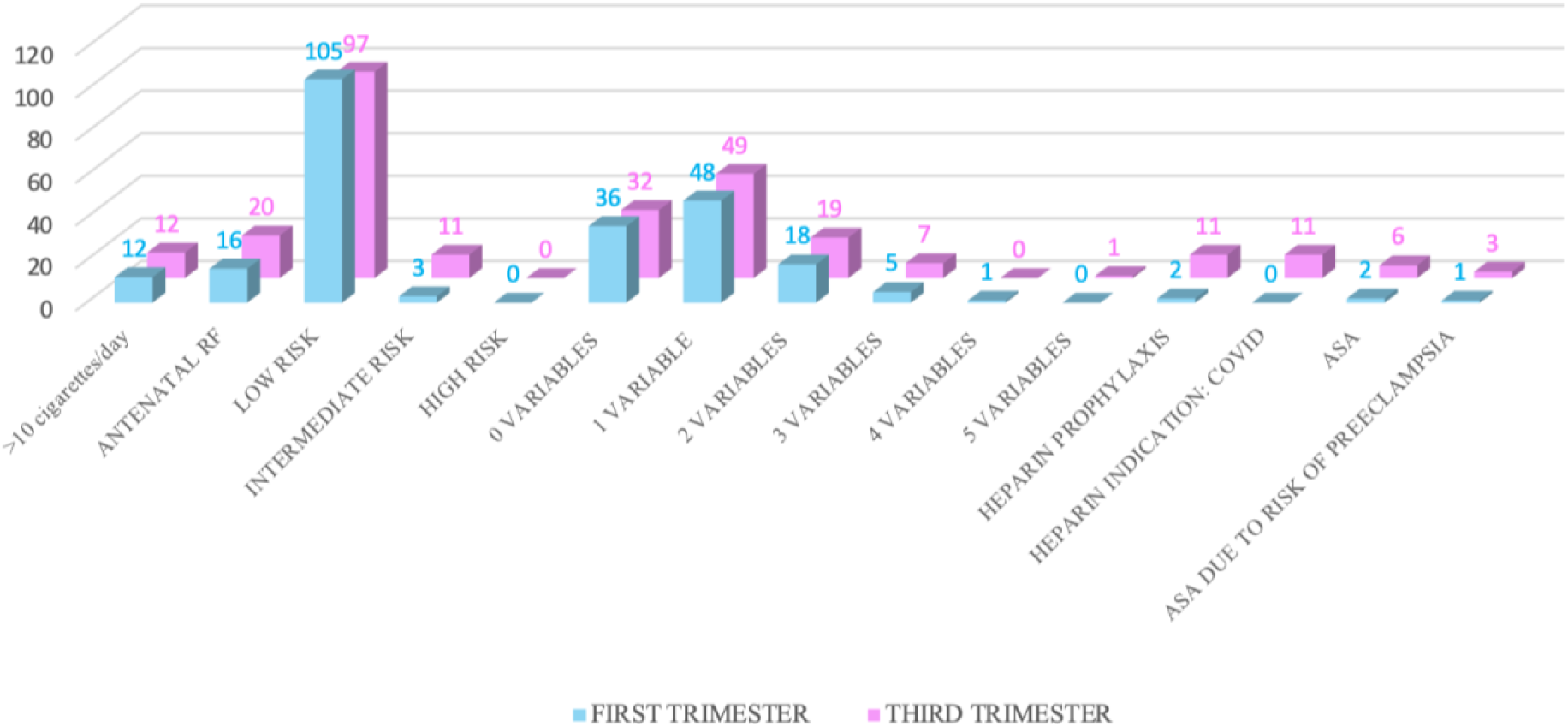
Comparison of risk factors between first and third trimester. Variation in the number of cigarettes consumed per day; antenatal risk factors; the number of patients with low, intermediate and high risk; the number of participants with positive variables in low risk (0, 1, 2, 3, 4 or 5); the number of pregnant women with LMWH prophylaxis and how many of these women had indication for it due to COVID-19; the number of patients with prescription of ASA and how many of these patients had indication for ASA due to risk of preeclampsia.

As for the number of pregnant women with low risk, in the first trimester it was 97.2%, while in the third trimester it was 89.8%. The number of patients with intermediate risk was 2.8% in the first trimester, but rises to 10.2% in the third trimester. However, the number of patients at high risk is null in both the first and third trimester.

In the first trimester, 33.3% of the patients did not present any positive variable at low risk, in contrast to the third trimester in which this percentage was 29.7%. In the first trimester, 44.4% of the patients had one positive variable at low risk, while in the third trimester this value was 45.4%. The percentage of patients with two positive variables in the first trimester was 16.7% and 17.6% in the third trimester. The percentage of pregnant women with three positive variables at low risk in the first trimester was 4.6%, in contrast to 6.5% in the third trimester. In the first trimester, 0.9% of the patients presented four risk variables and in the third trimester there were no patients with four variables. The percentage of pregnant women with five variables was 0% in the first trimester and 0.9% in the third trimester.

The number of pregnant women with heparin prophylaxis was of 2 patients in the first trimester, of which one was due to risk of VTE and previous miscarriages and another due to risk of VTE, previous miscarriages and IVF, and none due to COVID-19. This number rose to 11 in the third trimester, with 100% of the indications for SARS-CoV-2, and of these, 9.1% were also indicated for VTE risk, previous miscarriage and IVF.

There were 2 patients with prescription of ASA in the first trimester of gestation, of whom 50% had an indication for this drug due to risk of preeclampsia, and the other 50% due to family history. However, in the third trimester, 6 patients had an indication for ASA: 50% had a risk of preeclampsia, 16.7% had a prescription due to repeated miscarriages and another 16.7% due to family history. The remaining percentage had no clear indication.

### Types of delivery

Figure 5 shows the proportion of each type of delivery. Of the total 108 deliveries, 61 were eutocic (natural deliveries), 27 were performed by cesarean section (13 of which were cesarean sections decided during labour and 14 were programmed before the delivery) and 20 were performed with rotator instruments.

**Figure 5.**
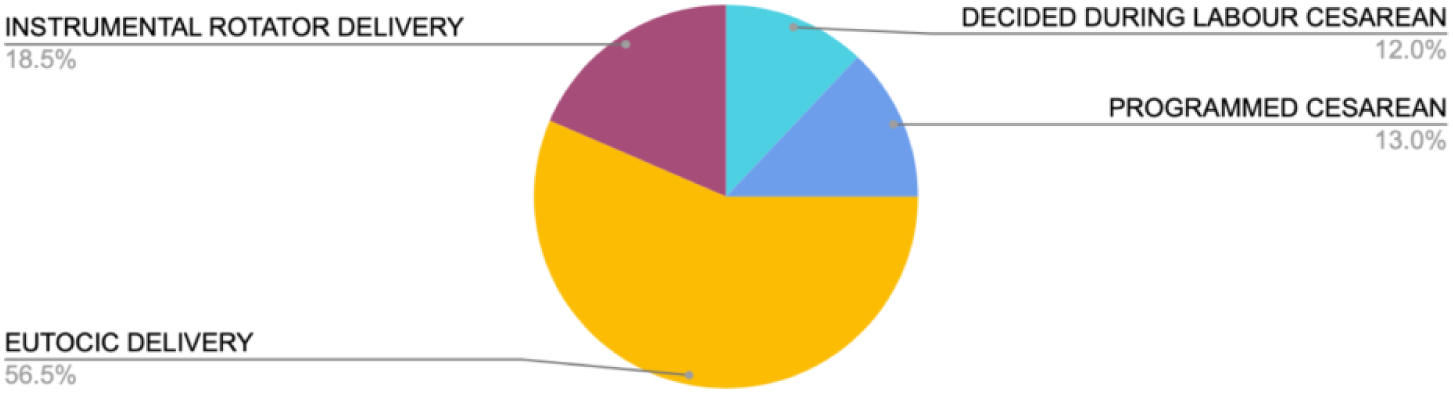
Types of delivery. Pie chart showing the proportion of each type of delivery: eutocic (natural childbirth), cesarean section scheduled or decided during labor, and instrumental rotator delivery

Figure 6 shows the parity of the women in the series (number of deliveries, including the one studied). Of the 108 patients, the delivery studied was the first in 51.9% of the women. In 36.1% of the women this delivery was the second, although one of them was primigravidae but she delivered twins, in 6.5% of the patients it was their third delivery and in 5.6% of the women it was the fourth.

**Figure 6.**
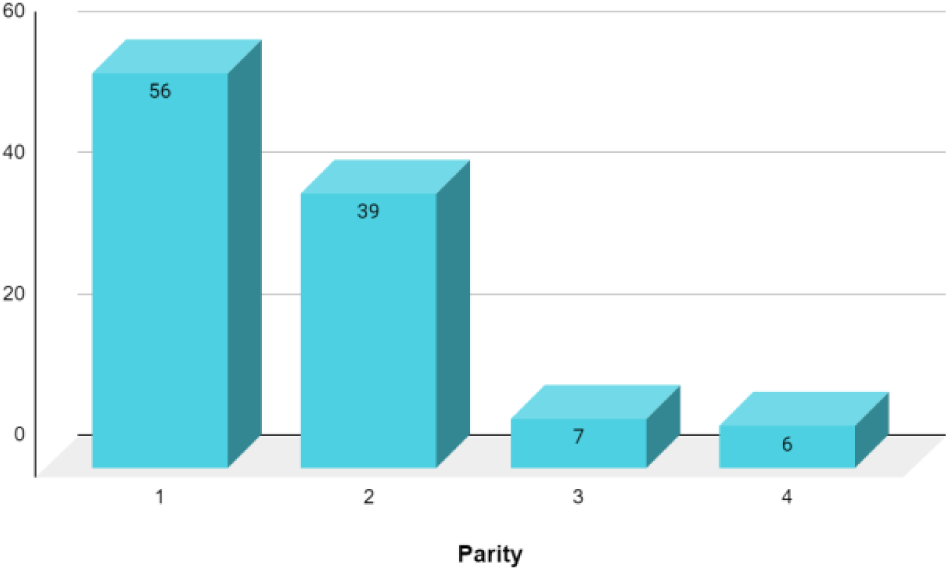
Parity (number of living children). The X-axis shows the parity of the patients (number of living children) and the Y-axis shows the number of women.

Regarding gestational age (the number of weeks of the baby born), it was observed that 9.3% of the deliveries were preterm (less than 37 weeks) and the rest were at term.

### Postpartum risk and heparin

Figure 7 shows the postpartum risk level of the different patients studied and the prescription of heparin according to that risk.

**Figure 7.**
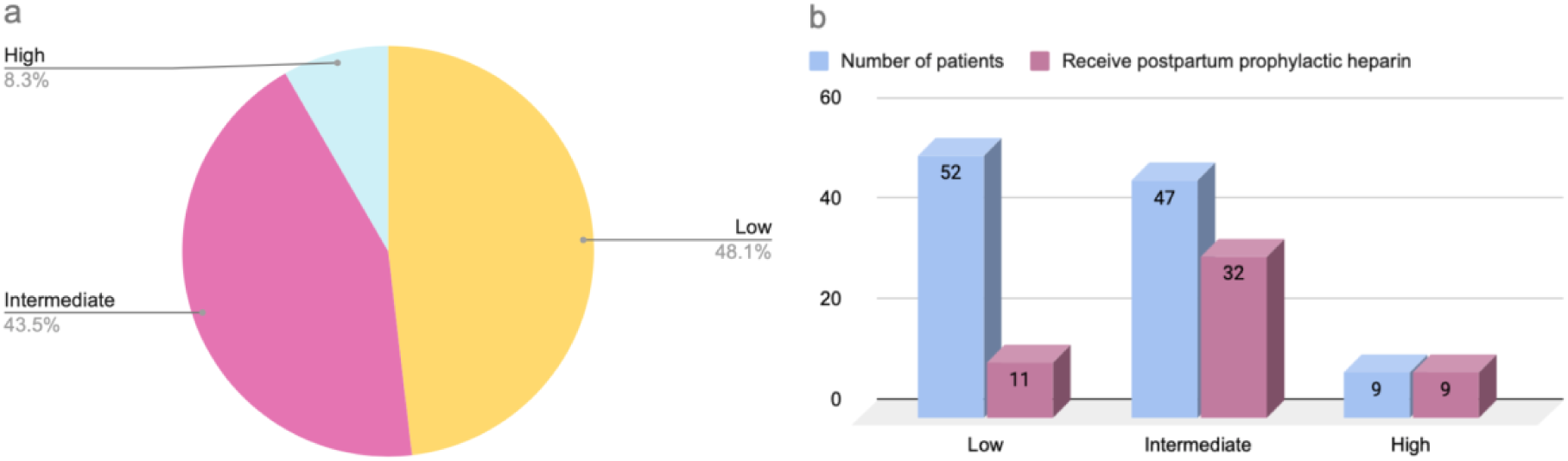
Risk factors and heparin. Postpartum risk level of the different patients studied. These patients have been assigned to levels based on the number of risk factors they present for VTE. Subsequently, the risk levels are related to prophylactic heparin treatment. Figure 7a shows the percentage of patients belonging to each risk level (high, intermediate or low). Figure 7b relates the number of patients in the different VTE risk levels (high, intermediate or low) to the number of patients treated with LMWH within each group.

Figure 7a shows the percentage of patients belonging to each risk level: high, intermediate or low. Of the total number of patients, 8.3% were at high risk of postpartum VTE, compared to a 43.5% who had intermediate risk. A total of 48.1% were at low risk of postpartum VTE. Figure 7b relates the number of patients at different levels of risk of VTE to the number of patients treated with LMWH within each group. Heparin was prescribed to 100% of patients at high risk of VTE in the postpartum period. Of the 47 patients belonging to the intermediate risk level, 68.1% were given prophylactic heparin. In addition, of the 52 patients assigned to a low VTE risk level, only 21.2% were treated with LMWH.

Furthermore, 48.1% of the total study participants were treated with LMWH during postpartum (all of them with enoxaparin), of whom 17.3% were at high risk of VTE, 61.5% at intermediate risk and 21.2% at low risk of VTE.

## Discussion

After following all the methodological steps of a research study, the risk factors of 108 women were analyzed longitudinally from the first trimester to the puerperium following uniform criteria. There are no studies in Spain that have longitudinally followed the risk factors in such depth throughout the entire pregnancy. This contrasts with the great relevance of LMWH during this period in the life of many women. Despite being a pilot study and, therefore, not having a large number of subjects, data and percentages similar to those of the Spanish population were obtained. Furthermore, the loss rate was only 4.4%.

It is noteworthy that 39.8% of the women were primigravidae. In addition, 11.1% smoked during pregnancy. The most relevant aspect of the family and personal history is the overall increase of 13 points in patients with COVID-19 due to the wave of the omicron variant that temporarily coincided with the third trimester of gestation of the women in the series. This is reflected in an increase in the prescription of LMWH in the third trimester. However, there is not a change in the baseline risk since there were no clear criteria and the *SARS-CoV-2* infection was not considered a risk variable.

There was a drop of 49 points in the number of patients at low risk from the first trimester to the puerperium, due to the addition of patients who gave birth by cesarean section to those who were already at risk during pregnancy. The cesarean section rate obtained is 25%, much higher than the 15% recommended by the World Health Organization (WHO). This value is not concordant with the average for this hospital, which was last calculated in 2019, obtaining a value of 19.8%. However, it does coincide with the Spanish average estimated by the *Instituto Nacional de Estadística* (INE) corresponding to 25%.

LMWH is a very common medication in pregnant women, with 53.6% of them requiring at least 10-14 days of heparin. The prescription rate in the HUPHM is in accordance with the Royal College guidelines 37a. The percentage of LMWH prescription is so high due to the high age of the pregnant women (38% over 35 years of age), a high rate of cesarean sections (25%), a high *SARS-CoV-2* infection (17.6%) and an IVF rate of 11.1%. All this requires that patients receive information, training and that they are discharged with the prescribed drug for self-administration.

The relevance of our study lies in the fact that 80.6% of the patients had at least one risk factor in the postpartum period and, from the second onwards, the prescription of heparin is recommended. Given that 34.3% of women had a risk factor, it is important to look for another possible factor involving the administration of LMWH. The importance of this relies on the fact that pulmonary thromboembolism is responsible for 20-30% of maternal deaths, being the seventh cause of mortality in these women (5).

This study was conducted at a particular time in Spanish obstetric history with one of the peaks of the COVID-19 pandemic that affected a huge number of pregnant women between December 2021 and January 2022. According to the *Centro de Coordinación de Alertas y Emergencias* of the Ministry of Health, the 14-day incidence of *SARS-CoV-2* infection averaged from December 2021 and January 2022 was of 2327.61. (6)

The percentage of patients with COVID-19 was 17.6%, but only 53% of these received LMWH. This reflects an evident heterogeneity in the criteria for prescribing this medication due to a lack of studies supporting prophylaxis in women infected with *SARS-CoV-2*. All this leads professionals to position themselves in two groups: a conservative one that does not administer the drug because there is no consensus on the scientific evidence and, another group that, empirically, decides to prescribe this medication seeking a balance between benefit and risk.

Given that the study is a pilot trial, it would be necessary to replicate the results with a larger number of participants.

## Conclusions

80.6% of the women in the study had some risk factor, therefore, the determination of these factors is critical. In conclusion, an assessment of thrombotic risk factors should be made in all pregnant women at the beginning of pregnancy, and should be repeated if any change in the variables is produced, as well as at the time of delivery and postpartum.

Prophylaxis with LMWH in pregnant women is very common in Spain, especially in the puerperium. This is due to the obstetric and demographic characteristics and, exceptionally, to the COVID-19 pandemic.

Finally, the heterogeneity in the prescription of LMWH in pregnant women infected with SARS-CoV-2 highlights the clear need for the development of clear protocols.

## Data Availability

All data produced in the present study are available upon reasonable request to the authors

## Abbreviations

ASA: acetylsalicylic acid
VTE: venous thromboembolic disease
IVF: in vitro fertilization
LMWH: low molecular weight heparins
HUPHM: *Hospital Universitario Puerta de Hierro Majadahonda*
BMI: body mass index
INE: *Instituto Nacional de Estadística*
WHO: World Health Organization,

## Acknowledgments

We would like to thank Dr. Hernández Guijo who, beyond his work as a tutor, gave us enough autonomy to make decisions according to our preferences without ceasing to be a point of reference. We would like to thank Dr. Martínez Pérez for his clinical support, his enthusiasm and dedication to research, as well as his commitment to our training. We cannot forget to thank all the participants and their families for their time, kindness and good disposition in such a special moment of life.

## References

(1) ACOG Practice Bulletin No. 196: Thromboembolism in Pregnancy. Obstet Gynecol 2018 -07;132(1):e1–e17.

(2) Rath W, Tsikouras P, von Tempelhoff G-. [Pharmacological Thromboprophylaxis during Pregnancy and the Puerperium: Recommendations from Current Guidelines and their Critical Comparison]. Z Geburtshilfe Neonatol 2016 -06;220(3):95–105.

(3) https://www.rcog.org.uk/guidance/browse-all-guidance/green-top-guidelines/reducing-the-risk-of-thrombosis-and-embolism-during-pregnancy-and-the-puerperium-green-top-guideline-no-37a/-Últimaentrada: 30.03.2022

(4) http://www.leo-pharma.es/Profesional-Sanitario/Trombosis/Calculadora-de-riesgo-ETV-OBS-App.aspx Última entrada: 25/03/2022

(5) Gallo-Vallejo JL, Naveiro-Fuentes M, Puertas-Prieto A, et al. Prevención del tromboembolismo venoso durante el embarazo y el puerperio en Atención Primaria y Especializada. Semergen. 2017;43(6):450–6. https://www.elsevier.es/es-revista-medicina-familia-semergen-40-articulo-prevencion-del-tromboembolismo-venoso-durante-S1138359316301988

(6) https://www.sanidad.gob.es/profesionales/saludPublica/ccayes/alertasActual/nCov/documentos/Actualizacion_533_COVID-19.pdf Last report entry for the month of December: 30/03/2022 https://www.sanidad.gob.es/profesionales/saludPublica/ccayes/alertasActual/nCov/documentos/Actualizacion_553_COVID-19.pdf Last report entry for the month of January: 30/03/2022

